# The Utility of ChatGPT as an Example of Large Language Models in Healthcare Education, Research and Practice: Systematic Review on the Future Perspectives and Potential Limitations

**DOI:** 10.1101/2023.02.19.23286155

**Authors:** Malik Sallam

## Abstract

An artificial intelligence (AI)-based conversational large language model (LLM) was launched in November 2022 namely, “ChatGPT”. Despite the wide array of potential applications of LLMs in healthcare education, research and practice, several valid concerns were raised. The current systematic review aimed to investigate the possible utility of ChatGPT and to highlight its limitations in healthcare education, research and practice. Using the PRIMSA guidelines, a systematic search was conducted to retrieve English records in PubMed/MEDLINE and Google Scholar under the term “ChatGPT”. Eligibility criteria included the published research or preprints of any type that discussed ChatGPT in the context of healthcare education, research and practice. A total of 280 records were identified, and following full screening, a total of 60 records were eligible for inclusion. Benefits/applications of ChatGPT were cited in 51/60 (85.0%) records with the most common being the utility in scientific writing followed by benefits in healthcare research (efficient analysis of massive datasets, code generation and rapid concise literature reviews besides utility in drug discovery and development). Benefits in healthcare practice included cost saving, documentation, personalized medicine and improved health literacy. Concerns/possible risks of ChatGPT use were expressed in 58/60 (96.7%) records with the most common being the ethical issues including the risk of bias, plagiarism, copyright issues, transparency issues, legal issues, lack of originality, incorrect responses, limited knowledge, and inaccurate citations. Despite the promising applications of ChatGPT which can result in paradigm shifts in healthcare education, research and practice, the embrace of this application should be done with extreme caution. Specific applications of ChatGPT in health education include the promising utility in personalized learning tools and shift towards more focus on critical thinking and problem-based learning. In healthcare practice, ChatGPT can be valuable for streamlining the workflow and refining personalized medicine. Saving time for the focus on experimental design and enhancing research equity and versatility are the benefits in scientific research. Regarding authorship in scientific articles, as it currently stands, ChatGPT does not qualify to be listed as an author unless the ICMJE/COPE guidelines are revised and amended. An initiative involving all stakeholders involved in healthcare education, research and practice is urgently needed to set a code of ethics and conduct on the responsible practices involving ChatGPT among other LLMs.

## 1. Introduction

Artificial intelligence (AI) can be defined as the multidisciplinary approach of computer science and linguistics that aspires to create machines capable of performing tasks that normally require human intelligence [1]. These tasks include the ability to learn, adapt, rationalize, understand and to fathom abstract concepts, as well as the reactivity to complex human attributes such as attention, emotion, creativity, etc. [2].

The history of AI as a scientific discipline can be traced back to the mid-XX century at the Dartmouth Summer Research Project on AI [3]. This was followed by the development of machine learning (ML) algorithms that allow decision making or predictions based on the patterns in large datasets [4]. Subsequently, the development of neural networks (brain mimicking algorithms), genetic algorithms (finding optimal solutions for complex problems by application of the evolutionary principles), and other advanced techniques followed [5].

Launched in November 2022, “ChatGPT” is an AI-based large language model (LLM) trained on massive text datasets in multiple languages with the ability to generate human-like responses to text input [6]. Developed by OpenAI (OpenAI, L.L.C., San Francisco, CA, USA), ChatGPT etymology is related to being a chatbot (a program able to understand and generate responses using a text-based interface) and being based on the Generative Pre-trained Transformer (GPT) architecture [6,7]. The GPT architecture utilizes a neural network to process natural language; thus, generating responses based on the context of input text [7]. The superiority of ChatGPT compared to its GPT-based predecessors can be linked to its ability to respond to multiple languages generating refined and highly sophisticated responses based on advanced modelling [6,7].

In the scientific community and academia, ChatGPT received mixed responses reflecting the history of controversy regarding the benefits vs. risks of advanced AI-technologies [8–10]. On one hand, ChatGPT among other LLMs, can be beneficial in conversational and writing tasks assisting to increase the efficiency and accuracy of the required output [11]. On the other hand, concerns raised in relation to possible bias based on the datasets used in ChatGPT training, which can limit its capabilities that could result in factual inaccuracies, yet alarmingly appearing scientifically plausible (a phenomenon termed hallucination) [11]. Additionally, security concerns and the potential of cyber-attacks with spread of misinformation utilizing LLMs should be considered as well [11].

The innate resistance of the human mind to any change is a well-described phenomenon and can be understandable from evolutionary and social psychology perspectives [12]. Therefore, the concerns and debates that arose instantaneously following the widespread release of ChatGPT appears understandable. The attention that ChatGPT received involved several disciplines. In education as an example, ChatGPT release could mark the end of essays as assignments [13]. In healthcare practice and academic writing, factual inaccuracies, ethical issues and fear of misuse including the spread of misinformation should be considered [14–16].

The versatility of human intelligence (HI) in comparison to AI should not be overlooked including its biologic evolutionary history, adaptability, creativity and the ability of emotional intelligence and to understand complex abstract concepts [2]. However, the HI-AI cooperation can have great benefits if accurate and reliable output of AI is ensured. The utility of AI in healthcare appears promising with possible applications in personalized medicine, drug discovery, and analysis of large datasets has been outlined previously besides the potential benefits in improving diagnosis and clinical decisions [17,18]. Additionally, an interesting area to probe is the utility of AI in healthcare education considering the massive information and various concepts that healthcare students have to grasp [19]. However, all these applications should be considered cautiously considering the valid concerns, risks and categorical failures experienced and cited in the context of LLM applications.

Therefore, the aim of the current review was to explore the future perspectives of ChatGPT as a prime example of LLMs in healthcare education, academic/scientific writing and healthcare practice based on the existing evidence. Importantly, the current review objectives extended to involve the identification of potential limitations and concerns associated with the application of ChatGPT in the aforementioned areas.

## 2. Materials and Methods

The current systematic review was conducted according to the Preferred Reporting Items for Systematic Reviews and Meta-Analyses (PRIMSA) guidelines [20].

The eligibility criteria involved any type of published scientific research or preprints (article, review, communication, editorial, opinion, etc.), addressing ChatGPT that fell under the following categories: (1) healthcare practice/research; (2) healthcare education; and (3) academic writing.

The exclusion criteria included: (1) non-English records; (2) records addressing ChatGPT in subjects other than those mentioned in the eligibility criteria; and (3) articles from non-academic sources (e.g., newspapers, internet websites, magazines, etc.).

The information sources included PubMed/MEDLINE and Google Scholar.

The exact PubMed/MEDLINE search strategy was as follows: (ChatGPT) AND ((“2022/11/30”[Date - Publication] : “3000”[Date - Publication])) which yielded 42 records. The search concluded on 16 February 2023.

The search on Google Scholar was conducted using Publish or Perish (Version 8) [21]. The search term was “ChatGPT” for the years: 2022–2023. The Google Scholar search yielded 238 records and concluded on 16 February 2023.

Then, the results from both searches were imported to EndNote v.20 for Windows (Thomson ResearchSoft, Stanford, CA, USA), which yielded a total of 280 records.

Then, screening of the title/abstract was done and duplicate records were excluded (*n* = 40), followed by exclusion of records published in languages other than English (*n* = 32). Additionally, the records that fell outside the scope of the review (records addressing ChatGPT in a context outside healthcare education, healthcare practice or scientific research/academic writing) were excluded (*n* = 80). Moreover, the records published in non-academic sources (e.g., newspapers, magazines, internet websites, blogs, etc.) were excluded (*n* = 18).

Afterwards, full screening of the remaining records (*n* = 110) was done. Screening completed which resulted in the exclusion of an additional 41 records that fell outside the scope of the current review. An additional nine records were excluded due to inability to access the full text being a subscription-based record.

This yielded a total of 60 records being eligible for inclusion in the current review.

Each of the included records were searched specifically for the following: (1) type of record (preprint, published research article, opinion, commentary, editorial, review, etc.); (2) the listed benefits/applications of ChatGPT in healthcare education, healthcare practice or scientific research/academic writing; (3) the listed risks/concerns of ChatGPT in healthcare education, healthcare practice or scientific research/academic writing; and (4) the main conclusions and recommendation regarding ChatGPT in healthcare education, healthcare practice or scientific research/academic writing.

Categorization of the benefits/applications of ChatGPT was as follows: (1) educational benefits in healthcare education (e.g., generation of realistic and variable clinical vignettes, customized clinical cases with immediate feedback based on the student’s needs, enhanced communications skills); (2) benefits in academic/scientific writing (e.g., text generation, summarization, translation and literature review in scientific research); (3) benefits in scientific research (e.g., efficient analysis of large datasets, drug discovery, identification of potential drug targets, generation of codes in scientific research); (4) benefits in healthcare practice (e.g., improvements in personalized medicine, diagnosis, treatment, life style recommendations based on personalized traits, documentation/generation of reports); and (5) being a freely available package.

Categorization of the risks/concerns of ChatGPT was as follows: (1) ethical issues (e.g., risk of bias, discrimination based on the quality of training data, plagiarism); (2) hallucination (the generation of scientifically incorrect content that sounds plausible); (3) transparency issues (black box application); (4) risk of declining need for human expertise with subsequent psychologic, economic and social issues; (5) over-detailed, redundant, excessive content; (6) concerns about data privacy for medical information; (7) risk of declining clinical skills, critical thinking and problem-solving abilities; (8) legal issues (e.g., copyright issues, authorship status); (9) interpretability issues; (10) referencing issues; (11) risk of academic fraud in research; (12) incorrect content; and (13) infodemic risk

## 3. Results

A total of 280 records were identified, and following the full screening process, a total of 60 records eligible to be included in the review. The PRISMA flow chart of the record selection process is shown in (Figure 1).

**Figure 1.**
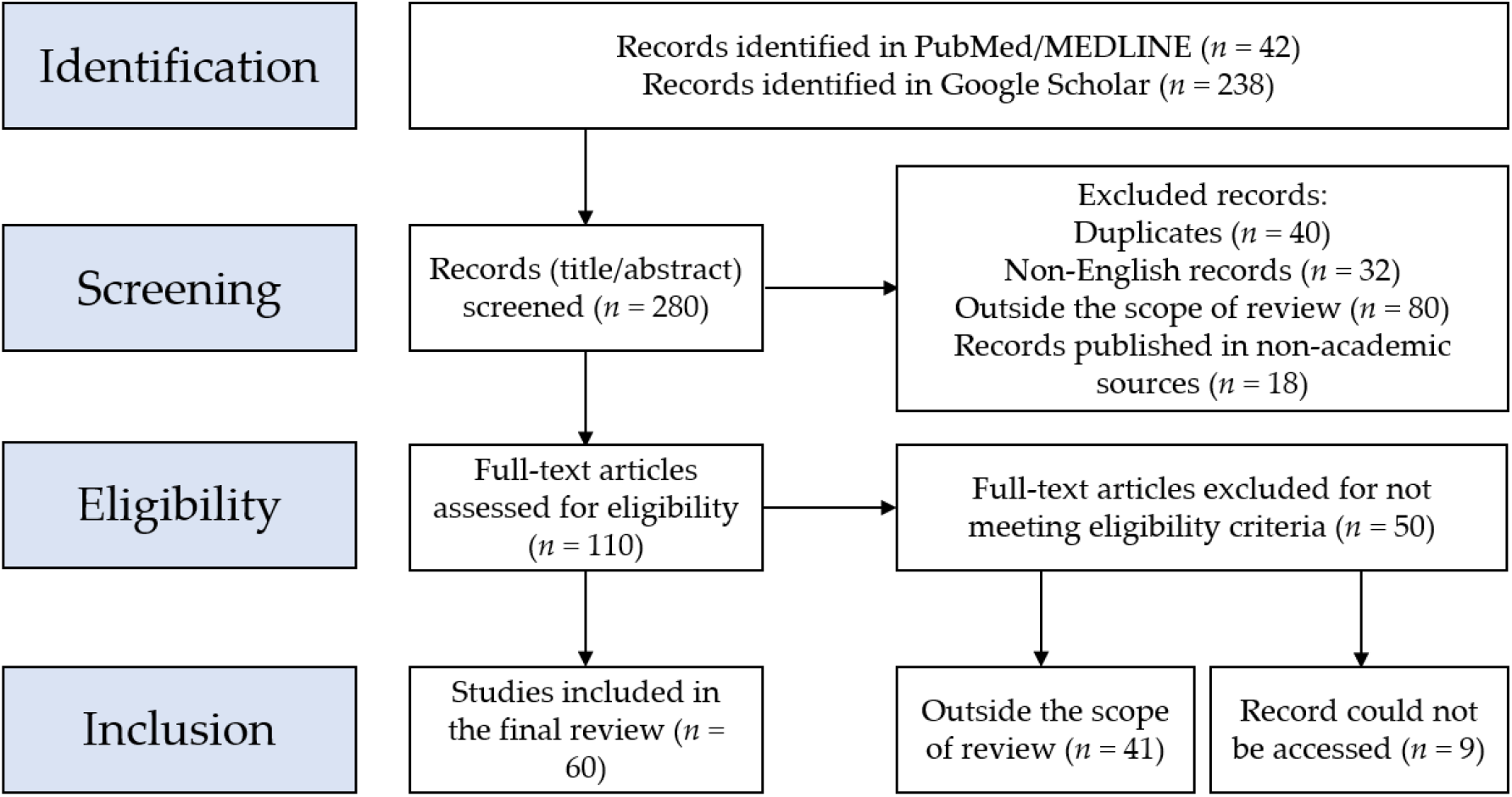
Flow chart of the record selection process based on the Preferred Reporting Items for Systematic Reviews and Meta-Analyses (PRIMSA) guidelines.

### 3.1. Summary of the ChatGPT Benefits and Limitations/Concerns in Healthcare

A summary of the main conclusions of the included studies regarding ChatGPT utility in academic writing, healthcare education and healthcare practice/research is provided in (Table 1).

**Table 1.** A summary of the main conclusions of the included studies.

| Author(s)<br>[Record] | Design, aims | Applications, benefits | Risks, concerns, limitations | Suggested action, conclusions |
| --- | --- | --- | --- | --- |
| Stokel-Walker<br>[13] | News explainer | Well-organized content with decent references;<br>free package | Imminent end of conventional educational<br>assessment; concerns regarding the effect on<br>human knowledge and ability | Revising educational assessment to prioritize<br>critical thinking or reasoning |
| Kumar [22] | Brief report; assessment of<br>ChatGPT for academic<br>writing in biomedicine | Original, precise and accurate responses with<br>systematic approach; helpful for training and to<br>improving topic clarity; efficiency in time;<br>promoting motivation to write | Instances of failure to follow the instructions<br>correctly; failure to cite references in-text<br>inaccurate references; lack of practical examples;<br>lack of personal experience highlights; superficial<br>responses | ChatGPT can help in improving academic writing<br>skills; promote universal design for learning;<br>proper use of ChatGPT under academic<br>mentoring |
| Wang et al. [23] | arXiv preprint;<br>investigating ChatGPT<br>effectiveness to generate<br>Boolean queries for<br>systematic literature<br>reviews | Higher precision compared to the current<br>automatic query formulation methods | Possibly not suitable for high-recall retrieval;<br>many incorrect MeSH terms; variability in query<br>effectiveness across multiple requests; a black-box<br>application | A promising tool for research |
| Borji [24] | arXiv preprint; to highlight<br>the limitations of ChatGPT | Extremely helpful in scientific writing | Problems in spatial, temporal, physical,<br>psychological and logical reasoning; limited<br>capability to calculate mathematical expressions;<br>factual errors; risk of bias and discrimination;<br>difficulty in using idioms; lack of real emotions<br>and thoughts; no perspective for the subject; over-detailed; lacks human-like divergences; lack of<br>transparency and reliability; security concerns<br>with vulnerability to data poisoning; violation of<br>data privacy; plagiarism; impact on the<br>environment and climate; ethical and social<br>consequences | Implementation of responsible use and<br>precautions; proper monitoring; transparent<br>communication; regular inspection for biases,<br>misinformation, among other harmful purposes<br>(e.g., identity theft) |
| Zielinski et al.<br>[25] | WAME recommendations<br>on ChatGPT | Can be a useful tool for researchers | Incorrect or non-sensical answers; restricted<br>knowledge to the period before 2021; no legal<br>personality; plagiarism | ChatGPT does not meet ICMJE criteria and<br>cannot be listed as an author; authors should be<br>transparent regarding ChatGPT use and take<br>responsibility for its content; editors need<br>appropriate detection tools for ChatGPT-generated content |
| Chen [26] | Editorial on ChatGPT applications in scientific writing | It helps to overcome language barriers promoting equity in research | Ethical concerns (ghostwriting); doubtful accuracy; citation problems | Embrace this innovation with an open mind; authors should have proper knowledge on how to exploit AI tools |
| Biswas [27] | Perspective record on the future of medical writing in light of ChatGPT | Improved efficiency in medical writing | Suboptimal understanding of the medical field; ethical concerns; risk of bias; legal issues; transparency issues | A powerful tool in the medical field; however, its several limitations should be considered |
| Thorp [28] | Editorial: "ChatGPT is not an author" | - | Content is not original; incorrect answers that sound plausible; issues of referencing; plagiarism | Revise assignments in education<br>In *Science* journals, the use of ChatGPT is considered a scientific misconduct |
| Kitamura [29] | Editorial on ChatGPT and the future of medical writing | Improved efficiency in medical writing; translation | Ethical concerns, plagiarism; lack of originality; inaccurate content; risk of bias | "AI in the Loop: Humans in Charge" |
| Stokel-Walker [30] | News article on the view of ChatGPT as an author | - | Plagiarism; lack of accountability; concerns about misuse in the academia | ChatGPT is not an author |
| Lubowitz [31] | Editorial, ChatGPT impact on medical literature | - | Inaccuracy; bias; spread of misinformation and disinformation; lack of references; redundancy in text | Authors should not use ChatGPT to compose any part of scientific submission; however, it can be used under careful human supervision to ensure the integrity and originality of the scientific work |
| van Dis et al. [32] | Comment: Priorities for ChatGPT research | Accelerated innovation; increased efficiency in publication time; can make science more equitable; increase the diversity of scientific perspectives; more free time for experimental designs; it could optimize academic training | Compromise research quality; transparency issues; spread of misinformation; inaccuracies, bias and plagiarism; ethical concerns; possible future monopoly; lack of transparency | Banning ChatGPT will not work; develop rules for accountability, integrity, transparency and honesty; carefully consider which academic skills remain essential to researchers; widen the debate in the academia; an initiative is needed to address the development and responsible use of LLM for research |
| Lund and Wang [33] | News: ChatGPT impact in academia | Useful for literature review; data analysis; translation | Ethical concerns, issues about data privacy and security; bias; transparency issues | ChatGPT has the potential to advance academia; consider how to use ChatGPT responsibly and ethically |
| Cotton et al. [34] | EdArXiv preprint on the academic integrity in ChatGPT era | - | Plagiarism; academic dishonesty | Careful thinking of educational assessment tools |
| Gao et al. [35] | bioRxiv preprint comparing the scientific abstracts generated by ChatGPT to original abstracts | A tool to decrease the burden of writing and formatting; can help to overcome language barriers | Misuse to falsify research; bias | The use of ChatGPT in scientific writing or assistance should be clearly disclosed |
| Liebrezn et al. [36] | Comment on the ethical issues of ChatGPT use in medical publishing | Can help to overcome language barriers | Ethical issues (copyright, attribution, plagiarism, and authorship); inequalities in scholarly publishing; spread of misinformation; inaccuracy | Implement robust AI author guidelines in scholarly publishing; follow COPE AI in decision making; AI cannot be listed as an author and its use must be properly acknowledged |
| Polonsky and Rotman [37] | SSRN preprint on listing ChatGPT as an author | Accelerate the research process; increase accuracy and precision | Intellectual property issues if financial gains are encountered | AI can be listed as an author in some instances |
| Nature [38] | Nature editorial on the rules of ChatGPT use to ensure transparent science | Can summarize research papers; generate helpful computer code | Ethical issues; transparency | LLM tools will be accepted as authors; if LLM tools are to be used, it should be documented in the methods or acknowledgements; advocate for transparency in methods, and integrity and truth from researchers |
| Aczel and Wagenmakers [39] | PsyArXiv preprint as a guide of transparent ChatGPT use in scientific writing | - | Issues of originality, transparency | Provide sufficient information, accreditation and verification of ChatGPT use |
| Manohar and Prasad [40] | A case study written with ChatGPT assistance | Helped to generate a clear, comprehensible text | Lack of scientific accuracy and reliability; citation inaccuracy | ChatGPT use must be discouraged because it can provide false information and non-existent citations; can be misleading in healthcare practice |
| Akhter and Cooper [41] | A case study written with ChatGPT assistance | Helped provide a relevant general introductory summary | Inability to access relevant literature; the limited knowledge up to 2021; citation inaccuracy; limited ability to critically discuss results | Currently, ChatGPT does not replace independent literature reviews in scientific research |
| Holzinger et al. [42] | Article: AI/ChatGPT use in biotechnology | Biomedical image analysis; diagnostics and disease prediction; personalized medicine; drug discovery and development | Ethical and legal issues; limited data availability to train the models; reproducibility of the runs | Aspire for fairness, open science, and open data |
| Mann [43] | Perspective: ChatGPT in translational research | Efficiency in writing; analysis of large datasets (e.g., electronic health records or genomic data); predict risk factors for disease; predict disease outcomes | Quality of data available; inability to understand the complexity of biologic systems | ChatGPT role in scientific and medical journals will grow in the near future |
| De Angelis et al. [44] | SSRN preprint discussing the concerns of an AI-driven infodemic | Can support and expediting academic research | Generation of misinformation and the risk of subsequent infodemics; falsified or fake research; ethical concerns | Carefully weigh ChatGPT benefits vs. risks; establish ethical guidelines for use; encourage a science-driven debate |
| Benoit [45] | medRxiv preprint on the generation, revision, and evaluation of clinical vignettes as a tool in health education using ChatGPT | Consistency, rapidity and flexibility of text and style; ability to generate plagiarism-free text | Clinical vignettes' ownership issues; inaccurate or non-existent references | ChatGPT can allow for improved medical education; better patient communication |
| Sharma and Thakur [46] | ChemRxiv preprint on ChatGPT possible use in drug discovery | Identify and validate new drug targets; design new drugs; optimize drug properties; assess toxicity; generate drug-related reports | Reliance on the data available for training which can result in bias or inaccuracy; inability to understand the complexity of biologic systems; transparency issues; lack of experimental validation; limited interpretability; limited handling of uncertainty; ethical issues | ChatGPT can be a powerful and promising assisting in drug discovery; however, ethical issues should be addressed |
| Moons and Van Bulck [47] | Editorial on ChatGPT potential in cardiovascular nursing practice and research | Summarize a large text; facilitate the work of researchers; data collection | Information accuracy issues; the limited knowledge up to 2021; limited capacity | ChatGPT can be a valuable tool in healthcare |
| Patel and Lam [48] | Comment on ChatGPT utility in documentation of discharge summary | Can help to reduce the burden of discharge summaries providing high-quality and efficient output | Data governance issues; risk of depersonalization of care; risk of incorrect or inadequate information | Proactive adoption to limit future issues |
| Cahan and Treutlein [49] | Editorial reporting a conversation with ChatGPT on stem cell research | It saves time | Repetition; several responses lacked depth and insight; lack of references | ChatGPT helped to write an editorial saving time |
| Rao et al. [50] | medRxiv preprint on the usefulness of ChatGPT in radiologic decision making | Moderate accuracy to determine appropriate imaging steps in breast cancer screening and evaluation of breast pain | Lack of references; alignment with user intent; inaccurate information; over-detailed; recommending imaging in futile situations; providing rationale for incorrect imaging decisions; black box nature | Using ChatGPT for radiologic decision making is feasible, potentially improving the clinical workflow and responsible use of radiology services |
| Antaki et al. [51] | medRxiv preprint on assessing ChatGPT to answer a diverse MCQ exam in ophthalmology | Currently performing at the level of an average first-year ophthalmology resident | Inability to process images; risk of bias; dependence on training dataset quality | There is a potential of ChatGPT use in ophthalmology; however, its applications should be approached carefully |
| Ahn [52] | Letter to the editor reporting a conversation of ChatGPT regarding CPR | Personalized interaction; quick response time; can help to provide easily accessible and understandable information regarding CPR to the general public | Inaccurate information might be generated with serious medical consequences | Explore the potential utility of ChatGPT to provide information and education on CPR |
| Gunawan [53] | An editorial reporting a conversation with ChatGPT regarding the future of nursing | Increased efficiency; reduce errors in care delivery | Lack of emotional and personal support | ChatGPT can provide valuable perspectives in healthcare |
| D'Amico et al. [54] | Editorial reporting a conversation of ChatGPT regarding incorporating Chatbots into neurosurgical practice and research | Can help to provide timely and accurate information for the patients about their treatment and care | Possibility of inaccurate information; privacy concerns; ethical issues; legal issues; bias; | Neurosurgery practice can be leading in utilizing ChatGPT into patient care and research |
| Aydın and Karaarslan [55] | SSRN preprint on the use of ChatGPT to conduct a literature review on digital twin in healthcare | Low risk of plagiarism; accelerated literature review; more time for researchers | Lack of originality | Expression of knowledge can be accelerated using ChatGPT; further work will use ChatGPT in citation analysis to assess the attitude towards the findings |
| Zhavoronkov * [56] | Perspective reporting a conversation with ChatGPT about Rapamycin use from a philosophical perspective | Provided correct summary of rapamycin side effects. Referred to the need to consult a healthcare provider based on the specific situation | - | Demonstration ChatGPT potential to generate complex philosophical arguments |
| Hallsworth et al. [57] | A comprehensive opinion article submitted before ChatGPT launching on the value of theory-based research | Can help to circumvent language barriers; can robustly help to process massive data in short time; can spark creativity by humans if “AI in the Loop: Humans in Charge” is applied | Ethical issues; legal responsibility issues; lack of empathy and personal communication; lack of transparency | There is an intrinsic value of human engagement in the scientific process which cannot be replaced by AI contribution |
| Sanmarchi et al. [58] | medRxiv preprint evaluating ChatGPT support to conduct an epidemiologic study following the STROBE recommendations<br>conduction of an epidemiological study | Can provide appropriate responses if properly queried; more time for researchers to focus on experimental phase | Bias in the training data; devaluation of human expertise; risk of scientific fraud; legal issues; reproducibility issues | The research premise and originality will remain the function of human brain; however, ChatGPT can assist in reproducing the study |
| Stokel-Walker and Van Noorden [14] | Nature news feature article on ChatGPT implications in science | More productivity among researchers | Problems in reliability and factual inaccuracies; misleading information that seem plausible; over-detailed; bias; ethical issues; copyright issues | “AI in the Loop: Humans in Charge”; we are just at the beginning |
| Duong and Solomon [59] | medRxiv preprint evaluating ChatGPT vs. human responses to questions on genetics | Generation of rapid and accurate responses; easily accessible information for the patients with genetic disease and their families; can help can health professionals in the diagnosis and treatment of genetic diseases; Could make genetic information widely available and help non-experts to understand this information | Plausible explanations for incorrect answers; reproducibility issues | Value of ChatGPT use will increase in importance in research and clinical settings |
| Huh [60] | To compare ChatGPT performance on a parasitology exam to that of Korean medical students | Performance will improve by deep learning | ChatGPT performance was lower compared to medical students; Plausible explanations for incorrect answers | ChatGPT performance will continue to improve, and healthcare educators/students are advised on how to incorporate it into the education process |
| Yeo et al. [61] | medRxiv preprint evaluating ChatGPT responses to questions on cirrhosis and hepatocellular carcinoma | Improved health literacy with better patient outcome; free availability; increased efficiency among health providers; emulation of empathetic responses | Non-comprehensive responses; the limited knowledge up to 2021; responses can be limited and not tailored to specific country or region; legal issues | ChatGPT may serve as a useful adjunct tool for patients besides the standard of care; future studies are recommended |
| Bašić et al. [62] | arXiv preprint on the performance of ChatGPT in essay writing compared to masters forensic students in Croatia | - | Plagiarism; lack of originality; it did not accelerate essay writing | The concerns in the academia towards ChatGPT are not totally justified; ChatGPT text detectors can fail |
| Fijačko et al. [63] | Letter to the editor to report the accuracy of ChatGPT responses to life support exam questions by the AHA | Relevant, accurate responses on occasions | Referencing issues; over-detailed | ChatGPT did not pass any of the exams; however, it can be a powerful self-learning tool to prepare for the life support exams |
| Hisan and Amri [64] | RG preprint on ChatGPT use medical education | Generation of educational content; useful to learn languages | Ethical concerns; scientific fraud (papermills); inaccurate responses; declining quality of educations with the issues of cheating | Appropriate medical exam design, especially for practical skills |
| Jeblick et al. [65] | arXiv preprint on ChatGPT utility to simplify and summarize radiology reports | Generation of medical information relevant for the patients; moving towards patient-centered care; cost efficiency | Bias and fairness issues; misinterpretation of medical terms; imprecise responses; odd language; hallucination (plausible yet inaccurate response); unspecific location of injury/disease | Demonstration of the ability of ChatGPT simplified radiology reports; however, the limitations should be considered<br>Improvements of patient-centered care in radiology could be achieved |
| Mbakwe et al. [66] | Editorial on ChatGPT ability to pass the USMLE | - | Bias; lack of thoughtful reasoning | ChatGPT passing the USMLE revealed the deficiencies in medical education and assessment; there is a need to reevaluate medical student training and education |
| Khan et al. [67] | Communication on ChatGPT use in medical education and clinical management | Automated scoring; assistance in teaching; improved personalized learning; assistance in research; generation of clinical vignettes; rapid access to information; translation; documentation in clinical practice; support in clinical decisions; personalized medicine | Lack of human-like understanding; the limited knowledge up to 2021 | ChatGPT can be a helpful in medical education, research, and clinical practice; however, it cannot replace the human capabilities |
| Gilson et al. [68] | Performance of ChatGPT on USMLE | Ability to understand context and to complete a coherent and relevant conversation in the medical field; can be used as an adjunct in group learning | The limited knowledge up to 2021 | ChatGPT passes the USMLE with performance at a 3<sup>rd</sup> year medical student level; can help to facilitate learning as a virtual medical tutor |
| Nisar and Aslam [69] | SSRN preprint on the assessment of ChatGPT usefulness to study pharmacology | Good accuracy | Content not sufficient for research purposes | Can be a helpful self-learning tool |
| Huh [70] | Editorial of JEEHP policy towards ChatGPT use | - | Responses not accurate in some areas | JEEHP will not accept ChatGPT as an author; however, ChatGPT content can be used if properly cited and documented |
| O'Connor * [71] | Editorial written with ChatGPT assistance on ChatGPT in nursing education | Personalized learning experience | Plagiarism; biased or misleading results | Advocate ethical and responsible use of ChatGPT; improve assessment in nursing education |
| Kung et al. [72] | ChatGPT raised accuracy to enable passing the USMLE | Accuracy with high concordance and insight; facilitate patient communication; personalized medicine | - | ChatGPT can have promising potential in medical education; recommendation for future studies to consider non-biased approach with quantitative natural language processing and text mining tools such as word network analysis |
| Lin [73] | PsyArXiv preprint describing the utility of ChatGPT in academic education | Versatility | Hallucination (inaccurate information that sounds scientifically plausible); fraudulent research; plagiarism; copyright issues | Transforming long-term effects; embrace ChatGPT and use it to augment human capabilities; however, sensible guidelines and codes of conduct are urgently needed |
| Shen et al. [74] | Editorial on ChatGPT strengths and limitations | Generation of medical reports; providing summary of medical records; drafting a letter to the insurance provider; improve the interpretability of CAD systems | Hallucination (inaccurate information that sounds scientifically plausible); the need to carefully craft questions or prompts; possible inaccurate or incomplete results; dependence on the training data; bias; research fraud | Despite the extremely helpful powers of ChatGPT, we should proceed cautiously in harnessing its power |
| Gordijn and Have [75] | Editorial on the revolutionary nature of ChatGPT | - | Factual inaccuracies; plagiarism; fraud; copyright infringements | We should prepare for a future with LLM has the capacity to write papers that pass peer review |
| Mijwil et al. [76] | Editorial on the role of cybersecurity in the protection of medical information | Versatility; efficiency; high quality of text generated; cost saving; room for innovation; improved decision making; improved diagnostics; predictive modeling; personalized medicine; streamline clinical workflow increasing efficiency; remote monitoring | Data security issues | Emphasis on the role of cybersecurity to protect medical information |
| The Lancet Digital Health [77] | Editorial on the strengths and limitations of ChatGPT | Improve language and readability | Over-detailed; incorrect or biased content; potentially generating harmful errors; spread of misinformation; plagiarism; issues with integrity of scientific records | Widespread use of ChatGPT is inevitable; proper documentation of its use; ChatGPT should not be listed or cited as an author or co-author |
| Aljanabi et al. [78] | Editorial on the possibilities provided by ChatGPT | Assist in academic writing; code generation | Inaccurate content including inability to reliably handle mathematical calculations | ChatGPT will receive a growing interest in the scientific community |
| Marchandot et al. [79] | Commentary on ChatGPT in academic writing | Assist in literature review saving time; ability to summarize papers; improving language | Inaccurate content; bias; may lead to decreased critical thinking and creativity in science; ethical concerns; plagiarism | ChatGPT can be credited as an author based on its significant contribution |

### 3.2. Characteristics of the Included Records

A summary of the record types included in the current review is shown in (Figure 2).

**Figure 2.**
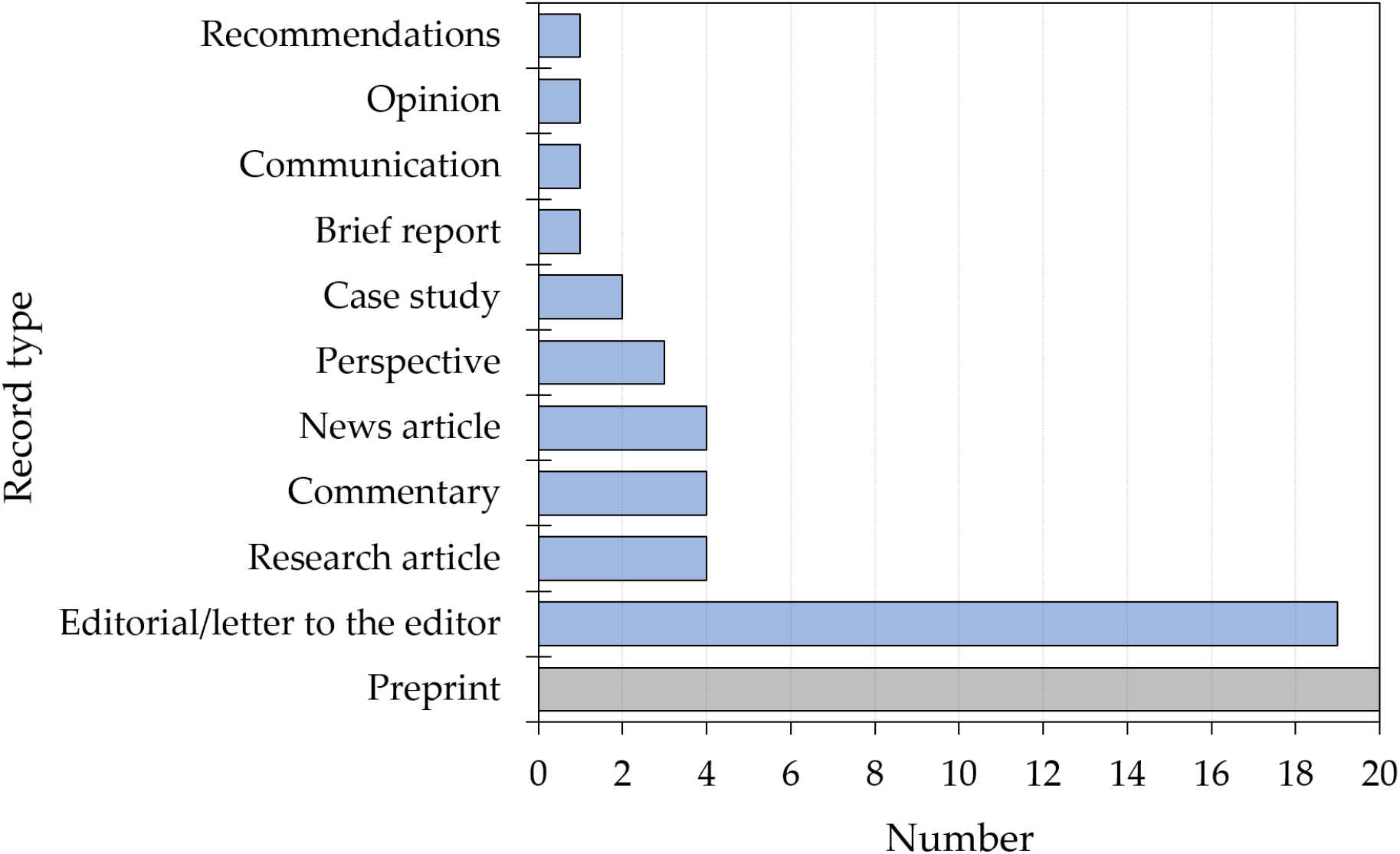
Summary of types of the included records (*n* = 60). Preprints (not peer reviewed) are highlighted in grey while published records are highlighted in blue.

One-third of the included records were preprints (*n* = 20), with the most common preprint server being medRxiv (*n* = 6, 30.0%), followed by SSRN and arXiv (*n* = 4, 20.0%) for each. Editorials/letters to editors were the second most common type of the included records (*n* = 19, 31.7%).

### 3.3. Benefits and Possible Applications of ChatGPT in Healthcare Education, Practice and Research Based on the Included Records

Benefits of ChatGPT was most frequently cited in the context of academic/scientific writing which was mentioned in 31 records (51.7%). Examples included: efficiency and versatility in writing with text of high quality, improved language, readability and translation promoting research equity, accelerated literature review. Benefits in scientific research followed which was mentioned in 20 records (33.3%). Examples included the ability to analyze massive data including electronic health records or genomic data, availability of more free time for the focus on experimental design, and drug design and discovery. Benefits in healthcare practice was mentioned by 14 record (23.3%), with examples including personalized medicine, prediction of disease risk and outcome, streamlining the clinical workflow, improved diagnostics, documentation, cost saving, and improved health literacy. Educational benefits in healthcare disciplines were mentioned in seven records (11.7%) with examples including: generation of accurate and versatile clinical vignettes, improved personalized learning experience, and being an adjunct in group learning. Being a free package was mentioned as a benefit in two records (3.3%, Figure 3).

**Figure 3.**
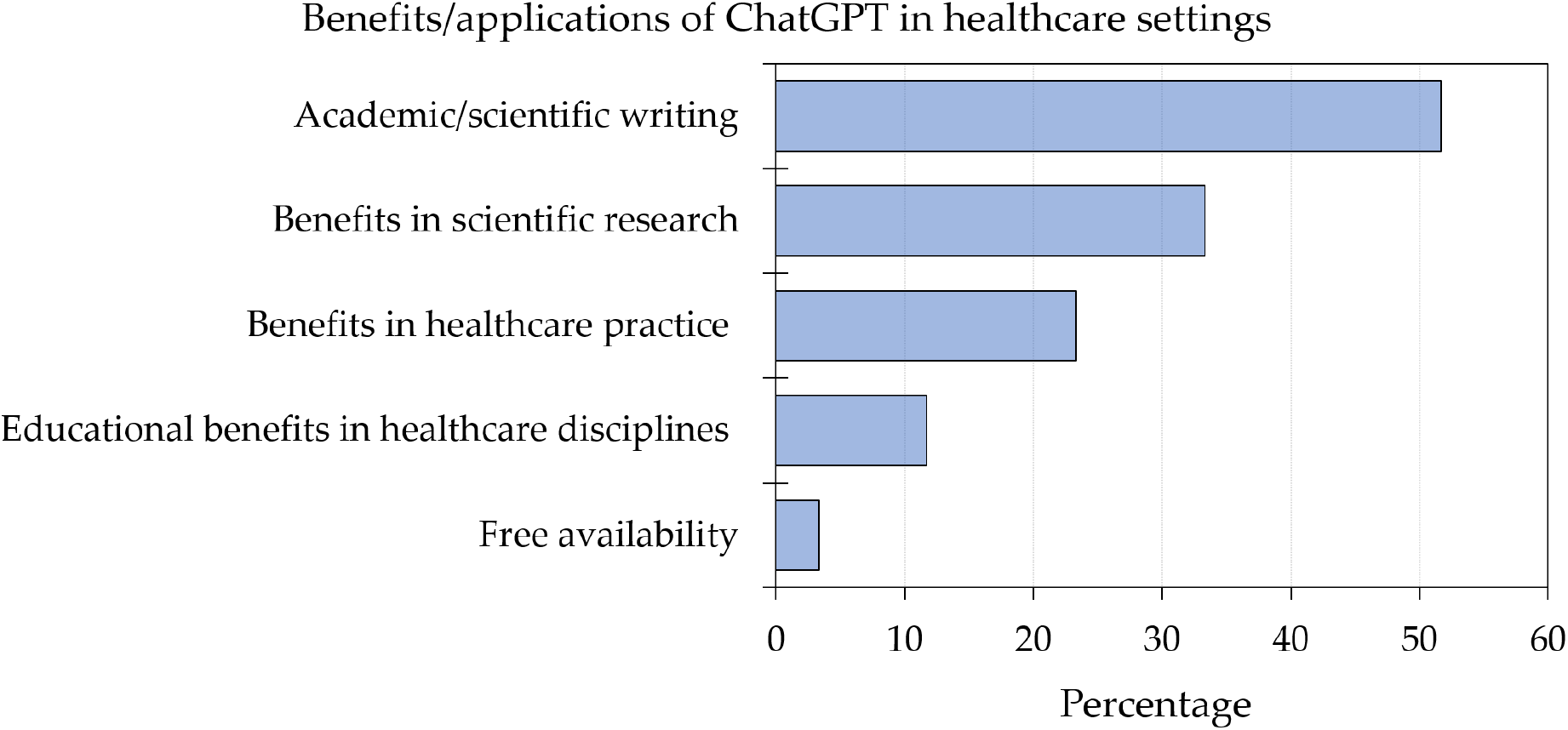
Summary of benefits/applications of ChatGPT in healthcare education, research and practice based on the included records.

### 3.4. Risks and Concerns Towards ChatGPT in Healthcare Education, Practice and Research Based on the Included Records

Ethical concerns were commonly mentioned by 33 records (55.0%), especially in the context of risk of bias (mentioned by 18 records, 30.0%) and plagiarism (mentioned by 14 records, 23.3%) among data privacy and security issues.

Other concerns involved: the risk of incorrect/inaccurate information that was mentioned by 20 records (33.3%); citation/reference inaccuracy or inadequate referencing which was mentioned by 10 records (16.7%); transparency issues which was mentioned by 10 records (16.7%); legal issues mentioned in 7 records (11.7%); restricted knowledge before 2021 mentioned by 6 records (10.0%); risk of misinformation spread mentioned by 5 records (8.3%); over-detailed content mentioned in 5 records (8.3%); copyright issues mentioned in 4 records (6.7%); and lack of originality mentioned by 4 records (6.7%, Figure 4).

**Figure 4.**
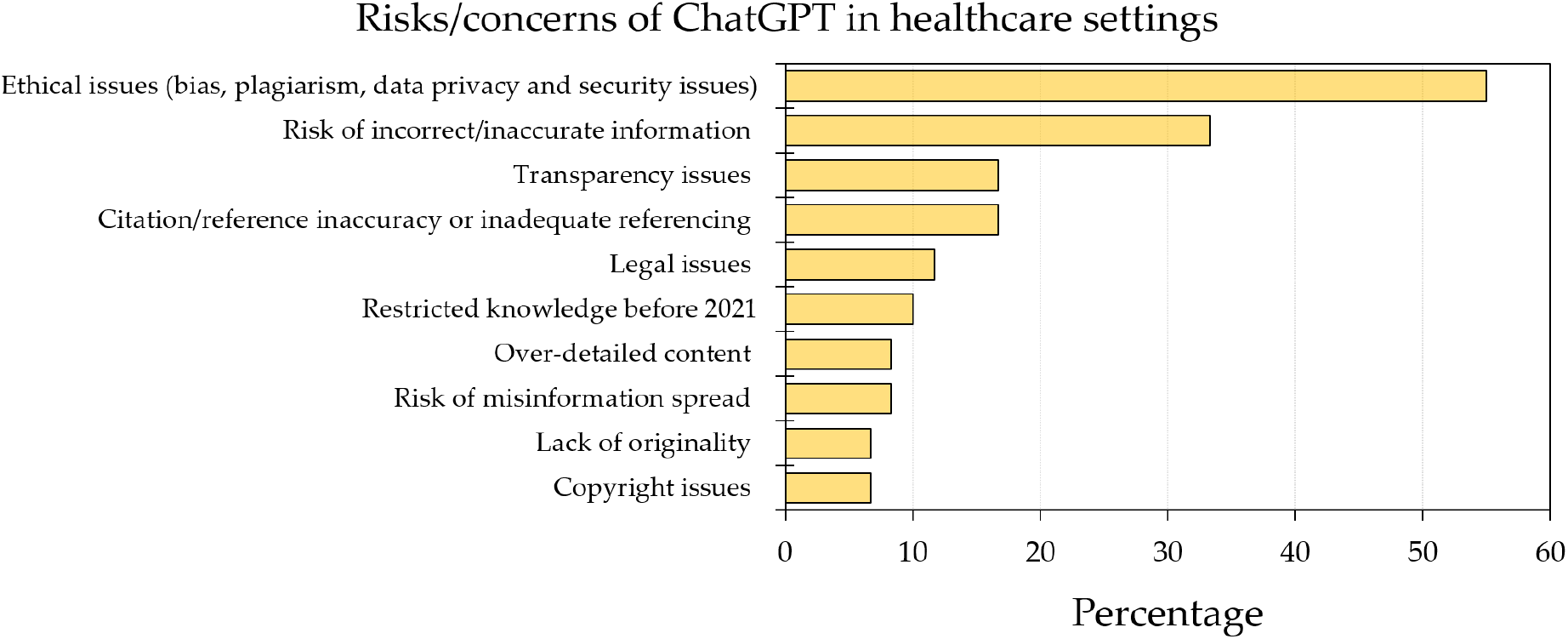
Summary of risks/concerns of ChatGPT use in healthcare education, research and practice based on the included records.

## 4. Discussion

The far-reaching consequences of ChatGPT among other LLMs can be described as a paradigm shift in the academia and healthcare practice [16]. The discussion of its potential benefits, future perspectives and importantly its limitations appear timely and relevant.

Therefore, the current review aimed to highlight these issues based on the current evidence. The following common themes emerged from the available literature: ChatGPT as an example of other LLMs can be a promising or even a revolutionary tool for scientific research both in academic writing and in the research process itself. Specifically, ChatGPT was listed in several sources as an efficient and promising tool for conducting comprehensive literature reviews and generating computer codes, thereby saving time for the research steps that require more efforts from human intelligence (e.g., the focus on experimental design) [14,27,32,33,38,47,55,58,78,79]. Additionally, ChatGPT can be helpful in generating queries for comprehensive systematic review with high precision as shown by Wang et al. despite the authors highlighting the transparency issues and unsuitability for high-recall retrieval [23]. Moreover, the utility of ChatGPT extends to involve the improvement in language and better ability to express and communicate the research ideas and results, ultimately speeding up the publication process with faster availability of research results [26,29,35,36,44,49,77]. This is particularly relevant for researchers who are non-native English speakers [26,29,35]. Such a practice can be acceptable considering the already existent English editing services provided by several academic publishers as an acceptable practice. Subsequently, this can help to promote equity and diversity in research [32,57].

On the other hand, the use of ChatGPT in academic writing and scientific research should be done in light of the following current limitations that could compromise the quality of research: First, superficial, inaccurate or incorrect content was frequently cited as a shortcoming of ChatGPT use [14,47,49,77,79]. The ethical issues including the risk of bias based on training datasets, and plagiarism were frequently mentioned, besides the lack of transparency described on occasions as a black box technology [14,27,29–33,35,36,39,57,58,77,79]. Importantly, the concept of ChatGPT hallucination was mentioned which can be risky if not evaluated properly by researchers and health providers with proper expertise [59,60,65,73,74]. This comes in light of ChatGPT’s ability to generate incorrect content that appears plausible scientifically.

Second, several records mentioned the current problems regarding citation inaccuracies, insufficient references and ChatGPT referencing to non-existent sources [26,31]. This was clearly shown in two recently published case studies with ChatGPT use in a journal contest [40,41,49]. These case studies discouraged the use of ChatGPT citing lack of scientific accuracy, limited updated knowledge, and lack of ability to critically discuss the results [40,41,75]. Therefore, the ChatGPT generated content albeit efficient should be meticulously examined prior to its inclusion in any research manuscripts or proposals for grants.

Third, the generation of non-original, over-detailed or excessive content can be an additional burden for the researchers who should carefully supervise ChatGPT-generated content [14,28,29,31,39,55]. This can be addressed by supplying ChatGPT with proper prompts (text input) due to varying responses based on the construction of prompts in order to be able to generate succinct content [58].

Fourth, as it currently stands, the knowledge of ChatGPT is limited to the period prior to 2021 based on the training datasets used in ChatGPT training [6]. Thus, ChatGPT currently cannot be used as a reliable updated source of literature review; nevertheless, it can be used as a motivation to organize the literature in a decent format and if supplemented by reliable and up-to-date references [47,61].

Fifth, the risk of research fraud (e.g., ghostwriting, falsified or fake research) involving ChatGPT should be considered seriously [26,44,58,73–75], as well as the risk of generating mis- or dis-information with subsequent possibility of infodemics [31,32,36,44].

Sixth, legal issues were raised by several records as well, including copyright issues [14,27,57,73,75]. Finally, the issue of listing ChatGPT as an author does not appear acceptable based on the current ICMJE and COPE guidelines for determining authorship as illustrated by Zielinski et al. and Liebrenz et al. [25,36]. This comes in light of the fact that authorship entails legal obligations which are not met by ChatGPT [25,36]. However, other researchers suggested the possibility of inclusion of ChatGPT as an author in some specified instances [37,79].

A few instances were encountered where ChatGPT was cited as an author which can point to the initial perplexity by publishers regarding the role of LLM including ChatGPT in research [56,71]. The disapproval of including ChatGPT or any other LLM in the list of authors was clearly explained in *Science, Nature*, and the *Lancet* editorials referring to its use as a scientific misconduct, and this view was echoed by many scientists [28,30,38,70,77]. In case of ChatGPT use in the research process, several records advocated the need for proper and concise disclosure and documentation of ChatGPT or LLM use in the methodology or acknowledgement sections [35,39,70]. A noteworthy and comprehensive record by Borji can be used as a categorical guide for the issues and concerns of ChatGPT use especially in scientific writing [24].

From the perspective of healthcare practice, it seems that there is a careful excitement vibe regarding ChatGPT applications. The utility of ChatGPT to streamline the clinical workflow appears promising with expected increased efficiency in healthcare delivery and saving costs [53,65,74,76]. This was illustrated recently by Patel and Lam highlighting the ability of ChatGPT to produce efficient discharge summaries, which can be valuable to reduce the burden of documentation in healthcare [48]. Additionally, ChatGPT among other LLMs can have a transforming potential in healthcare practice via enhancing diagnostics, prediction of disease risk and outcome, and drug discovery among other areas in translational research [42,43,46]. Moreover, ChatGPT showed moderate accuracy to determine the imaging steps needed in breast cancer screening and evaluation of breast pain which can be a promising application in decision making in radiology [50]. There is also the prospects of personalized medicine and improved health literacy by providing easily accessible and understandable health information for the general public [52,54,59,61,72]. This utility was demonstrated by ChatGPT responses highlighting the need to consult healthcare providers among other reliable sources on specific situations [16,56].

On the other hand, several concerns regarding ChatGPT use in healthcare settings were raised. Ethical issues including the risk of bias and transparency issues appear as a recurring major concern [42,46,50,65]. Generation of inaccurate content can have severe negative consequences in healthcare; therefore, this valid concern should be cautiously considered [48,52,54]. This can also extend to involve providing justification for incorrect decisions [50].

Issues of interpretability, reproducibility and handling of uncertainty were raised as well, which can have harmful consequences in healthcare settings including research [46,58,59]. In the area of personalized medicine, the transparency issues in terms of ChatGPT being a black box with unclear information regarding the source of data used for its training is an important issue considering the variability observed among different populations in several traits [50]. The issue of reproducibility between prompt runs is of particular importance in healthcare practice [42].

Medico-legal issues and accountability in case of medical errors caused by ChatGPT application should be carefully considered [27]. Importantly, the current LLMs including ChatGPT does not appear to be able to comprehend the complexity of biologic systems needed in healthcare decisions and research [43,46]. The concerns regarding data governance, cybersecurity of medical information and data privacy should draw specific attention in the discussion regarding the utility of LLMs in healthcare [48,54,76].

Other issues include the lack of personal and emotional perspectives, which were listed among other concerns of ChatGPT utility in healthcare delivery and research [52,57]. However, ChatGPT emulation of empathetic responses was reported in a preprint in the context of hepatic disease [61]. Additionally, the issue of devaluing the function of the human brain should not be overlooked; therefore, stressing on the indispensable human role in healthcare practice and research is important to address any psychologic, economic and social consequences [58].

In the area of healthcare education, ChatGPT appears to have a massive transformative potential. The need to rethink and revise the current assessment tools in healthcare education comes in light of ChatGPT’s ability to pass reputable exams (e.g., USMLE), and possibility of ChatGPT misuse that would result in increased academic dishonesty [28,34,64,66,68,72].

Specifically, in ophthalmology examination, Antaki et al. showed that ChatGPT currently performed at the level of an average first-year resident [51]. The focus on questions involving the assessment of critical and problem-based thinking appears of utmost value [66]. Additionally, the utility of ChatGPT in healthcare education can involve tailoring education based on the student needs with immediate feedback [32]. Interestingly, a recent preprint by Benoit showed the promising potential of ChatGPT in rapidly crafting consistent realistic clinical vignettes of variable complexities which can be a valuable educational source with lower costs [45]. Thus, ChatGPT can be useful in healthcare education including enhanced communication skills given proper academic mentoring [22,45,67]. However, the copyright issue in ChatGPT-generated clinical vignettes besides the issue of inaccurate references should be taken into account [45]. Additionally, ChatGPT availability can be considered as a motivation in healthcare education based on the personalized interaction it provides, enabling powerful self-learning as well as its utility as an adjunct in group learning [52,63,67,68,71].

Other concerns in the educational context include the concern regarding the quality of training datasets resulting in bias and inaccurate information limited to the period prior to the year 2021, and current inability to handle images, as well as the low performance in some topics (e.g., failure to pass a parasitology exam for Korean medical students) and the issue of possible plagiarism [51,60,62,63,67,68]. Despite being described by versatility in the context of academic education [73], the content of ChatGPT in research assignments was discouraged as well, being currently insufficient, biased or misleading [69,71].

### 4.1. Future Perspectives

As stated comprehensively in a commentary by van Dis et al., there is an urgent need to develop guidelines for ChatGPT use in scientific research taking into account the issues of accountability, integrity, transparency and honesty [32]. Thus, the application of ChatGPT to advance academia should be done ethically and responsibly taking into account the risks and concerns it entails [33].

More studies are needed to evaluate the LLMs’ content, including its potential impact to advance academia and science with particular focus on healthcare settings. In academic writing, a question would arise if the authors would prefer an AI-editor and an AI-reviewer considering the previous flaws in the editorial and peer review processes [80–82]. A similar question would arise in healthcare settings involving the preference personal and emotional support from healthcare providers rather than the potential efficiency of AI-based systems

### 4.2. Strengths and Limitations

The current review represents the first rapid and concise overview of ChatGPT utility in healthcare education, research and practice. However, the results of the current review should be viewed carefully in light of several shortcomings that included: (1) the quality of the included records can be variable compromising generalizability of the results; (2) the exclusion of non-English records might have resulted in selection bias; (3) the exclusion of several records that could not be accessed could have resulted in missing relevant data despite being small in number; (4) the inclusion of preprints that have not been peer reviewed yet might compromise the generalizability of the results as well.

## Data Availability

Data supporting this systematic review are available in the original publications, reports and pre-prints that were cited in the reference section. In addition, the analyzed data that were used during the current systematic review are available from the author on reasonable request.

## 5. Conclusions

The imminent dominant use of LLM technology including ChatGPT utility will be inevitable. Considering the valid concerns raised over its potential misuse, particularly in the areas of healthcare education, research and practice, appropriate guidelines and regulations are urgently needed following the involvement of all stakeholder to help ensuring the harnessing of the potential powers of ChatGPT and other LLMs safely and responsibly. Ethical concerns, transparency, and legal issues should be considered carefully and proactive embrace of this technology can limit the future complications. If properly addressed, these technologies can have the potential to expedite the research and innovation in healthcare and can aid to promote equity in research by overcoming the language barriers. Therefore, a science-driven debate regarding the pros and cons of ChatGPT is strongly recommended and the possible benefits should be weighed with the possible risks of misleading results and fraudulent research.

Healthcare professionals could be described based on the available evidence as carefully enthusiastic considering the huge potential of ChatGPT among other LLMs in clinical decision making and optimizing the clinical workflow.

“ChatGPT in the Loop: Humans in Charge” can be the proper motto based on the intrinsic value of human knowledge and expertise in healthcare research and practice [14,57], reminiscent of the relationship of the human character Cooper and robotic character TARS from interstellar.

However, before its widespread adoption, ChatGPT impact in real world setting from healthcare perspective should be done (e.g., using a risk-based approach) [83]. Based on the title of an important perspective article “AI in the hands of imperfect users” by Kostick-Quenet and Gerke [83], Ferrari F2004 (the highly successful formula 1 racing car) broke Formula 1 records in the hands of Michael Schumacher; however, in my own hands - as a humble researcher without expertise in Formula 1 driving-it will only break walls and get broken beyond repair as well.

## Declarations

### Supplementary Materials

None.

### Author Contributions

The author explicitly and clearly declares the sole role in the following: conceptualization, methodology, software, investigation, resources, writing—original draft preparation, writing—review and editing, and visualization. The author explicitly and clearly declares that neither ChatGPT nor any other LLMs were used to draft this manuscript.

### Funding

This research received no external funding.

### Institutional Review Board Statement

Not applicable.

### Informed Consent Statement

Not applicable.

### Data Availability Statement

Data supporting this systematic review are available in the original publications, reports and preprints that were cited in the reference section. In addition, the analyzed data that were used during the current systematic review are available from the author on reasonable request.

## Acknowledgments

None.

## Conflicts of Interest

The author declares no conflict of interest.

## Notes

### Competing Interest Statement

The authors have declared no competing interest.

